# Examining National Health Insurance Fund Members’ preferences and trade-offs for the attributes of contracted outpatient facilities in Kenya: a discrete choice experiment

**DOI:** 10.1101/2024.07.16.24310505

**Authors:** Jacob Kazungu, Edwine Barasa, Justice Nonvignon, Matthew Quaife

**Author notes:** Joint Senior Author.

## Abstract

Patient choice of health facilities is increasingly gaining recognition for potentially enhancing the attainment of health system goals globally. In Kenya, National Health Insurance Fund (NHIF) members are required to choose an NHIF-contracted outpatient facility before accessing care. Understanding their preferences could support resource allocation decisions, enhance the provision of patient-centered care, and deepen NHIF’s purchasing decisions. We employed a discrete choice experiment to examine NHIF members’ preferences for attributes of NHIF-contracted outpatient facilities in Kenya. We developed a d-efficient experimental design with six attributes, namely availability of drugs, distance from household to facility, waiting time at the facility until consultation, cleanliness of the facility, attitude of health worker, and cadre of health workers seen during consultation. Data were then collected from 402 NHIF members in six out of 47 counties. Choice data were analysed using panel mixed multinomial logit and latent class models. NHIF members preferred NHIF-contracted outpatient facilities that always had drugs [β=1.572], were closer to their households [β=-0.082], had shorter waiting times [β=-0.195], had respectful staff [β=1.249] and had either clinical officers [β=0.478] or medical doctors [β=1.525] for consultation. NHIF members indicated a willingness to accept travel 17.8km if drugs were always available, 17.7km to see a medical doctor for consultation, and 14.6km to see respectful health workers. Furthermore, NHIF members indicated a willingness to wait at a facility for 8.9 hours to ensure the availability of drugs, 8.8 hours to see a doctor for consultation, and 7.2 hours to see respectful health workers. Understanding NHIF member preferences and trade-offs can inform resource allocation at counties, service provision across providers, and purchasing decisions of purchasers such as the recently formed social health insurance authority in Kenya as a move towards UHC.

## Introduction

Kenya is among many low- and middle-income countries that have committed to achieving universal health coverage (UHC) by 2030 (1, 2). To accelerate progress towards this goal, the government of Kenya has prioritized purchasing reforms and identified social health insurance through the National Health Insurance Fund (NHIF) as the ‘vehicle’ to drive the UHC agenda (3).

Over the years, in a bid to transform the NHIF into a strategic purchaser of health services (3), several purchasing reforms have been implemented, including the introduction of outpatient cover and the empanelment and contracting of health providers (4). However, transforming the NHIF into a strategic purchaser requires a continuous pursuit of better ways to act on the strategic purchasing actions including provider identification and selection and understanding of the population’s needs, preferences, and values (5).

While the introduction of outpatient cover allowed NHIF members to choose their preferred NHIF-contracted outpatient facilities with an option to change providers every quarter, there is a dearth of evidence highlighting the health facility attributes influencing NHIF members’ choice of contracted outpatient facilities in Kenya. Understanding NHIF members’ preferences is essential to tailor services that are attractive to people and could encourage demand thus informing health provider identification and contracting decisions by the NHIF. Besides, understanding NHIF members’ preferences could offer insights into resource allocation at the national and county levels. For instance, evidence of trade-offs such as NHIF members’ willingness to travel further to get medicine can inform the prioritization of medicines or training health workers to be respectful as opposed to the construction of new ill-equipped facilities. We used a discrete choice experiment (DCE) to understand the preferences NHIF members have for the attributes of NHIF-contracted outpatient facilities in Kenya.

DCEs are stated preference elicitation methods where respondents are asked to choose between two or more competing hypothetical alternatives with the alternatives differing across a range of attribute levels (6–8). Particularly, DCEs are useful when the researcher aims to understand trade-offs that respondents are willing to accept to compensate for other attributes, for example, the distance they are willing to travel for facilities with preferred characteristics.

Increasingly, DCEs are being used to examine either patients’ or a population’s preferences for health facilities (9–13). Evidence from these studies has highlighted several attributes summarised as ‘structure’, ‘process’, and ‘outcome’ attributes that influence patients/population preferences for health facilities (12). For instance, waiting time has been reported as the most important attribute in studies conducted in Western Cape Province (9), England (14), Ethiopia (15), China (16), and Liberia (17). Besides, other attributes such as the cost of care (18, 19), distance to facilities (15), availability of medicines and medical equipment (9, 19, 20), and staff attitude (18, 20) have also been reported to influence the choice of a facility. Despite the burgeoning evidence, these studies were either conducted in high-income countries, did not focus on patients or populations covered by a social health insurance scheme, and/or importantly, were not conducted in Kenya and therefore did not examine preferences among NHIF members even though the NHIF will drive the UHC agenda in Kenya. Consequently, we conducted a DCE among NHIF members in Kenya to understand context-specific and policy-relevant attributes and trade-offs that can inform optimal provider identification and selection by NHIF, resource allocation by other purchasers such as county departments of health, and patient-centred service delivery by health providers.

## Materials and methods

### Study setting and design

Kenya runs a devolved governance system comprised of a national government and 47 semi-autonomous county governments (21). As a result, health is one of the devolved functions in Kenya where the Ministry of Health executes policy and regulatory functions whilst running the national referral facilities (Level 6 facilities) whereas counties manage primary (Levels 2 to 4 facilities) and secondary facilities (Level 5 facilities). While the health financing system is described in detail elsewhere (22), in summary, in 2018/19, the health system was financed through government tax (47.6%), donor funding (19.1%), health insurance premiums (6.7%), and out-of-pocket payments by households (26.6%).

Kenya has one of the oldest public health insurance schemes in Africa – the National Health Insurance Fund (NHIF) - formerly known as the National Hospital Insurance Fund – started in 1966. As of 2022, the NHIF covered 24% of the population (23). Despite the relatively low population coverage compared to other countries such as Ghana (58.2%), Rwanda (78.7%) and Gabon (40.8%) (24), over the years, the NHIF has undergone several reforms aiming to transform it into a strategic purchaser of healthcare services (3, 25, 26). In 2015, the NHIF introduced an outpatient cover where members were required to voluntarily select outpatient facilities to access outpatient care while providers were paid through capitation (4). The NHIF contracts both public and private providers (both faith-based and private-for-profit providers). While the choice of outpatient providers (single health facilities) is voluntary, members are required to have selected an outpatient provider before accessing care but have the opportunity to change providers quarterly.

### Survey development

The study followed the recommended good research practices checklist for conducting DCEs by the International Society for Pharmacoeconomics and Outcomes Research Conjoint Analysis Task Force (27, 28).

While there are several preference elicitation methods in health (6), the study employed a DCE to permit the quantification of preferences across multiple attributes, and the quantification of the relative importance of the attributes whilst examining trade-offs NHIF members would be willing to make when selecting contracted outpatient facilities in Kenya. Besides, as a methodology, DCEs have been implemented in Kenya in several other topics such as to examine the preferences of healthcare providers for capitation (29), community health volunteer incentives (30), women’s place of childbirth (31), and clinical officer job preference (32), and thus considered appropriate for its use in this study.

### Attributes and levels

Attributes and attribute levels for the study were developed following the four-stage process of raw data collection, data reduction, attribute and attribute-level dropping, and wording (33). Raw data collection and data reduction were conducted following a literature review (34) and focus group discussions with NHIF members (35). The identified attributes (6 broad attributes from the literature and 7 from the FGDs) and levels were then examined by the researchers and 7 attributes were then piloted across 38 NHIF members. Based on the findings from the pilot data analysis, a list of six attributes was selected (Table 1Table 1). One attribute (opening hours of the facility) was dropped because it did not influence participants’ choices in the pilot (Results from the pilot – Supplementary file 1) and was ranked last in the FGDs.

**Table 1:** NHIF-contracted outpatient facility attributes and levels.

| Attribute | Stated levels | Definition | Attribute Type |
| --- | --- | --- | --- |
| Availability of drugs | Not always available | Whether prescribed drugs were available for free at the facility | Categorical |
|  | Always available |  |  |
| Distance from household to the facility | 1KM | Distance from the household to the facility | Continuous |
|  | 3KM |  |  |
|  | 5KM |  |  |
| Waiting time at the facility for consultation | 0.5 hours (30min) | Waiting time at the facility until consultation | Continuous |
|  | 1 hour |  |  |
|  | 2 hours |  |  |
| Attitude of health worker | Health worker is harsh and abusive | This referred to the way a health worker speaks to NHIF members at the facility | Categorical |
|  | Health worker is respectful |  |  |
| Cleanliness of the facility | Facility (toilets-rooms-floors) Not always clean | Whether the toilets, consultation rooms and floors were always clean | Categorical |
|  | Facility (toilets-rooms-floors) Always clean |  |  |
| The cadre of health workers seen for consultation | Nurse | The cadre of health workers seen by an NHIF member during consultation. | Categorical |
|  | Clinical Officer |  |  |
|  | Medical Doctor |  |  |

The definitions of all attributes are provided in Table 1. Further to the definition of the attribute ‘cadre of staff seen during the consultation’, in Kenya, there are three main cadres of staff that can see patients during a consultation: nurses, clinical officers, and medical doctors. Nurses often run lower-level facilities (Level 2 – dispensaries) where they can see patients for consultation but also provide support care such as drug administration, patient care, and health promotion in higher-level facilities. However, clinical officers and medical doctors often practice in higher-level facilities from Levels 3 to 6. While there are similarities in roles, Table 2 summarizes the differences between nurses, clinical officers, and medical doctors in Kenya.

**Table 2:**
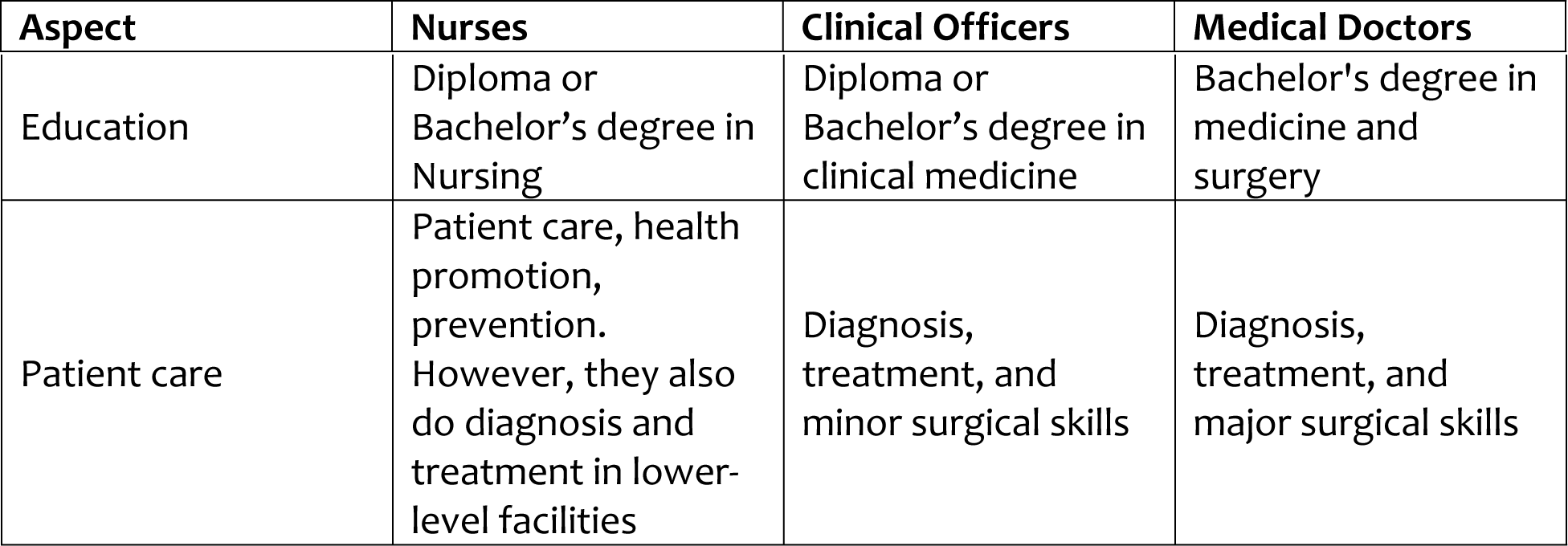
Summary of key differences between Nurses, Clinical Officers, and Medical Doctors.

| Aspect | Nurses | Clinical Officers | Medical Doctors |
| --- | --- | --- | --- |
| Education | Diploma or Bachelor's degree in Nursing | Diploma or Bachelor's degree in clinical medicine | Bachelor's degree in medicine and surgery |
| Patient care | Patient care, health promotion, prevention. However, they also do diagnosis and treatment in lower-level facilities | Diagnosis, treatment, and minor surgical skills | Diagnosis, treatment, and major surgical skills |

#### 1. Construction of choice tasks and experimental design

All six attributes (full profiles) were used to generate a fractional D-efficient experimental design in Ngene software 1.3.0 (36) using priors generated from the analysis of the pilot data (an orthogonal design used for the pilot and data analysed following a conditional logit model) – see Supplementary File 1 for priors obtained from the pilot. In this design, respondents were asked to choose between two unlabelled health facility alternatives (Health Facility A or Health Facility B) – Table 3. An opt-out option was not included in this design as it was deemed unrealistic as NHIF members must have chosen an outpatient facility before accessing care. In the choice tasks, we bolded some keywords in the levels to make them more visible for respondents to easily distinguish from the other levels of the attribute.

**Table 3:**
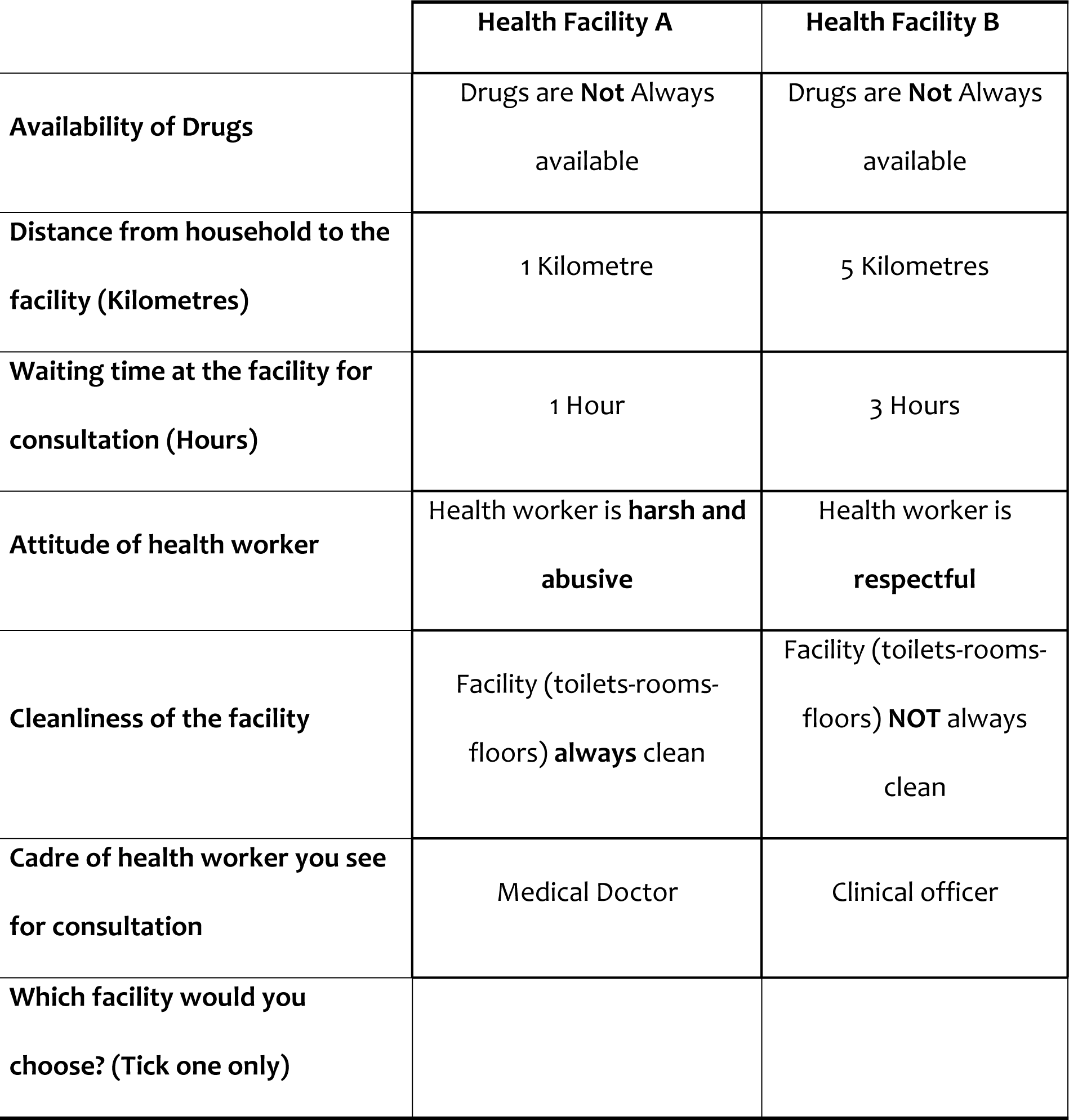
Sample choice task.

|  | **Health Facility A** | **Health Facility B** |
| --- | --- | --- |
| **Availability of Drugs** | Drugs are **Not** Always available | Drugs are **Not** Always available |
| **Distance from household to the facility (Kilometres)** | 1 Kilometre | 5 Kilometres |
| **Waiting time at the facility for consultation (Hours)** | 1 Hour | 3 Hours |
| **Attitude of health worker** | Health worker is **harsh and abusive** | Health worker is **respectful** |
| **Cleanliness of the facility** | Facility (toilets-rooms-floors) **always** clean | Facility (toilets-rooms-floors) **NOT** always clean |
| **Cadre of health worker you see for consultation** | Medical Doctor | Clinical officer |
| **Which facility would you choose? (Tick one only)** |  |  |

Given that having too many choice tasks has been associated with a cognitive burden on respondents (8, 37), our final design had 12 choice tasks. The 12 choice tasks were deemed appropriate as respondents had handled 12 choice tasks easily during the pilot and were within the number of tasks included in a majority of other studies (38, 39).

#### 2. Questionnaire development

After the final experimental design, we developed a paper-based questionnaire that contained five sections. Section A collected general information about the area where the survey was being conducted. Section B collected socio-demographic information about the respondents (including age, gender, and level of education). Section C explained the attributes and attribute levels while Section D depicted the 12 choice tasks. Section E asked for supplementary information about the respondents such as whether they had any chronic conditions or had already selected an NHIF-contracted outpatient facility.

#### 3. Sampling and data collection

Data for the DCE survey were collected across six counties: Kilifi, Taita Taveta, Makueni, Migori, Uasin Gishu, and Nyeri. These six counties were randomly selected from a list of all 47 counties in Kenya following a stratified simple random sampling approach without replacement (40). Prior to selection, counties were sorted from lowest to highest using the proportion of NHIF-accredited health facilities calculated as:

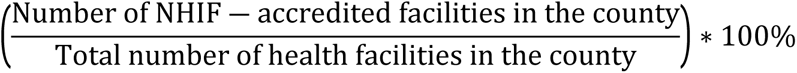

Counties were then stratified into three categories: 1) counties with a low proportion of NHIF-accredited facilities – operationally defined as counties with a proportion of less than 50% (a total of 17 counties); 2) counties with a medium proportion of NHIF-accredited facilities – operationally defined as counties whose proportion of NHIF accredited facilities was between 50% and less than 70% (a total of 16 counties; and 3) counties with a high proportion of NHIF-accredited facilities – operationally defined as counties with a proportion of 70% and above (a total of 14 counties). Two counties were then randomly selected within each stratum. The data for the total number of health facilities in each county and that of the number of NHIF-accredited facilities in each county was obtained from the Kenya Master Health Facility List (KMHFL) (41) and the list of health facilities offering the National Scheme was obtained from NHIF (42) respectively (Table 4).

**Table 4:** Selected County health facility statistics and sample sizes.

| County | Total Number of facilities in the county (A) | Total Number of NHIF-accredited facilities (B) | The proportion of outpatient NHIF-accredited facilities offering the National scheme (%) $[(B/A) * 100]$ | Target number of NHIF members (sample size) | The actual number of respondents |
| --- | --- | --- | --- | --- | --- |
| Makueni | 344 | 80 | 23% | 67 | 64 |
| Nyeri | 452 | 151 | 33% | 67 | 67 |
| Kilifi | 343 | 188 | 54% | 67 | 65 |
| Migori | 279 | 187 | 67% | 67 | 64 |
| Uasin Gishu | 230 | 161 | 70% | 67 | 68 |
| Taita Taveta | 113 | 81 | 72% | 67 | 70 |

From the S-error estimate of the D-efficient design, a minimum sample size of 310 respondents was estimated as required. However, to account for a potentially low response rate, uncertainty in the priors used from the pilot, and to allow proper distribution of respondents across the counties, we increased the minimum sample size by 30% resulting in a sample size of 403 which was distributed proportionally across the counties, thus, targeting 67 NHIF members in each of the counties. Each of the counties was then divided into rural and urban areas. Given challenges at the qualitative phase to randomly select participants from lists of NHIF members at NHIF (35), we used community health volunteers/promoters across rural and urban settings in each of the counties, to purposefully mobilize groups of 10-15 NHIF members from households to convene at a central location either in the rural or urban areas to complete the questionnaire. Data collectors guided the respondents throughout the entire process of completing the questionnaire. They ensured that each question was clearly understood by the respondents. Data collection was conducted between 21/11/2022 and 24/02/2023.

### Statistical analyses

The analysis of DCE data is grounded in the random utility theory (RUT) framework (8) and until recently, the random regret minimization theory (RRM) (43). The RUT assumes individuals are utility maximizers when selecting competing alternatives. Following Lancaster’s theory of consumer choice, individuals derive utility not from the goods or services themselves but from the attributes of the goods or services (44). Consequently, the utility an individual *i* derives from an alternative *j* in a choice scenario *s* can be broken down into a systematic component (specified as a function of the attributes of the alternative) *V_ijs_*and a random (unexplainable) component (representing unmeasured variations in preferences) *ε_ijs_* (8, 27).

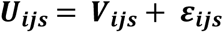

In the analysis, all attributes were dummy-coded except distance and waiting time which were continuous in kilometers and hours respectively. Based on the attributes presented in Table 1, the utility function of our DCE was defined as below:

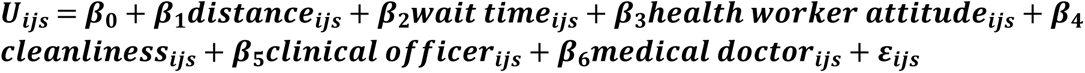

Several models were fitted in this analysis. First, we fitted a panel mixed multinomial logit model (MMNL) to account for preference heterogeneity between respondents whilst relaxing the independent from irrelevant alternatives (IIA) assumption under a conditional logit (MNL) model (7, 45). In this model, all parameters were assumed to be randomly distributed.

Second, we estimated the relative importance of the attributes following an approach described elsewhere (46). Essentially, using absolute coefficients from an MNL model, we 1) calculated the maximum effect by taking the product of the attribute coefficient and the largest difference between the attribute levels of the attribute, 2) summed up all the maximum effect values, 3) calculated the relative importance by dividing the maximum effect (computed in 1) by the sum of the maximum effects (computed in 2).

Third, following the panel MMNL model above, we computed two marginal willingness to accept (WTA) estimates in willingness to pay (WTP) space. Calculating WTA in WTP space has been shown to result in realistic values as opposed to estimating them in preference space (47), avoiding the generation of a Cauchy distribution as the ratio of two randomly distributed variables. In this study, we used distance and waiting time and estimated respondents’ willingness to accept travelling (WTT) to a facility and willingness to wait (WTW) in queue until getting a consultation. The *mixlogitwtp* command was used to compute the marginal WTA estimates in WTP space in Stata version 16.1 (48, 49).

Fourth, we fitted a latent class (LC) model with fixed parameters to explore unobserved heterogeneity (50). LC models assume that there are segments (classes) of the respondents where preferences are homogeneous within the classes but heterogeneous between the classes. We used the Akaike information criteria (AIC) and Bayesian Information Criteria (BIC) to select the optimal number of classes by comparing models with two to ten classes (49). The *lclogit* command was used to fit the LC model.

### Ethics statement

Ethical approval for the study was obtained from the Scientific Ethics Review Unit (SERU) of KEMRI (Ref: KEMRI/SERU/CGMR-C/191/4019). We obtained permissions in the form of letters of support or stamps from County Departments of Health in all study counties that facilitated entry into the county and facilities. Also, we obtained permission to conduct the study from the NHIF, the National Commission for Science, Technology, and Innovation (NACOSTI), and the Council of Governors in Kenya. Participants were taken through a consent process before completing the study questionnaire and once they understood the process of their involvement, they signed a consent form.

## Results

### Descriptive analysis

A majority of the NHIF members engaged in the survey were male (52.2% [95% CI: 47.3 – 57.1]), with a median age of 37 years[Inter-quartile range (IQR): 30 – 49], employed in the informal sector (36.1% [95% CI: 31.5 – 40.9]), liked that they were let to choose their NHIF contracted outpatient facilities (95.3% [95% CI: 92.7 – 97.0), and had already selected an NHIF-contracted outpatient facility (86.8% [95% CI: 83.1 – 89.8]) (Table 5).

**Table 5:** Characteristics of NHIF members engaged in the DCE.

|  | n | % | 95% Confidence Intervals |  |
| --- | --- | --- | --- | --- |
|  |  |  | Lower | Upper |
| **Gender** |  |  |  |  |
| Female | 192 | 47.8 | 42.9 | 52.7 |
| Male | 210 | 52.2 | 47.3 | 57.1 |
| **County** |  |  |  |  |
| Kilifi | 65 | 16.2 | 12.9 | 20.1 |
| Migori | 68 | 16.9 | 13.6 | 20.9 |
| Makueni | 64 | 15.9 | 12.7 | 19.8 |
| Nyeri | 67 | 16.7 | 13.3 | 20.6 |
| Uasin Gishu | 68 | 16.9 | 13.6 | 20.9 |
| Taita Taveta | 70 | 17.4 | 14.0 | 21.4 |
| **Age** |  |  |  |  |
| Mean | 402 | 40.0 | 38.7 | 41.3 |
| Median (*Inter-quartile range*) | 402 | 37.0 | 30.0 | 49.0 |
| **Employment status** |  |  |  |  |
| Not employed | 136 | 33.8 | 29.4 | 38.6 |
| Employed in the informal sector | 145 | 36.1 | 31.5 | 40.9 |
| Employed in the formal sector | 121 | 30.1 | 25.8 | 34.8 |
| **Distance to nearest contracted NHIF facility (KM)** |  |  |  |  |
| Mean (95% Confidence Interval) | 402 | 9.5 | 8.2 | 10.9 |
| Median (*Inter-quartile range*) | 402 | 5.0 | 2.0 | 11.0 |
| **Mode of transport to nearest NHIF-contracted outpatient facility** |  |  |  |  |
| Walking | 95 | 23.6 | 19.7 | 28.0 |
| Boda Boda (Motorbike) | 170 | 42.3 | 37.5 | 47.2 |
| Tuktuk | 4 | 1.0 | 0.4 | 2.6 |
| Public Transport/Matatu or Bus | 127 | 31.6 | 27.2 | 36.3 |
| Private car | 6 | 1.5 | 0.7 | 3.3 |
| **Whether respondent likes that they can choose an outpatient facility** |  |  |  |  |
| No | 19 | 4.7 | 3.0 | 7.3 |
| Yes | 383 | 95.3 | 92.7 | 97.0 |
| **Whether the respondent had chosen an NHIF-contracted outpatient facility** |  |  |  |  |
| No | 53 | 13.2 | 10.2 | 16.9 |
| Yes | 349 | 86.8 | 83.1 | 89.8 |

For the respondents that had already selected an NHIF-contracted outpatient facility (n=349), they were nearly equally distributed by gender, chose facilities where drugs were always available, lived on an average of 8.7 Kilometres (95% CI: 7.4 – 10.0) from their selected NHIF-contracted outpatient facility, and used Boda Boda (motorcycle) when traveling to their selected facility. On average, NHIF members waited at the facility for 1.3 hours (95% CI: 1.2 – 1.5), had selected facilities where health workers were respectful and saw a medical doctor during a consultation (Table 6).

**Table 6:** Socio-demographic and health facility factors for NHIF members that had selected an NHIF-contracted outpatient facility.

|  | n | Percentage | 95% Confidence Intervals |  |
| --- | --- | --- | --- | --- |
|  |  |  | Lower | Upper |
| Gender |  |  |  |  |
| Female | 173 | 49.6 | 44.3 | 54.8 |
| Male | 176 | 50.4 | 45.2 | 55.7 |
| Availability of Drugs at the selected facility |  |  |  |  |
| Not always available | 140 | 40.1 | 35.1 | 45.4 |
| Always available | 209 | 59.9 | 54.6 | 64.9 |
| **Distance from household to selected facility (KM)** |  |  |  |  |
| Mean (95% Confidence Interval) | 349 | 8.7 | 7.4 | 10.0 |
| Median (Inter-quartile range) | 349 | 5.0 | 2.0 | 10.0 |
| **Travel time to the selected facility** |  |  |  |  |
| Mean (95% Confidence Interval) | 349 | 1.1 | 0.8 | 1.5 |
| Median (Inter-quartile range) | 349 | 0.6 | 0.5 | 1.0 |
| **Means of transport used to travel to the facility** |  |  |  |  |
| Walking | 75 | 21.5 | 17.5 | 26.1 |
| Boda Boda (Motorcycle) | 175 | 50.1 | 44.9 | 55.4 |
| Tuktuk | 9 | 2.6 | 1.3 | 4.9 |
| Public Transport | 85 | 24.4 | 20.1 | 29.2 |
| Private car | 5 | 1.4 | 0.6 | 3.4 |
| **Waiting time at the facility until consultation** |  |  |  |  |
| Mean (95% Confidence Interval) | 349 | 1.3 | 1.2 | 1.5 |
| Median (Inter-quartile range) | 349 | 1.0 | 0.5 | 2.0 |
| **Attitude of health workers at the chosen facility** |  |  |  |  |
| Health worker is harsh and abusive | 19 | 5.4 | 3.5 | 8.4 |
| Health worker is respectful | 330 | 94.6 | 91.6 | 96.5 |
| **Cleanliness of the chosen facility** |  |  |  |  |
| Facility (toilets-rooms-floors) are not always clean | 41 | 11.8 | 8.8 | 15.6 |
| Facility (toilets-rooms-floors) are always clean | 307 | 88.2 | 84.4 | 91.2 |
| **Cadre of health worker seen at the facility during a consultation** |  |  |  |  |
| Nurse | 41 | 11.8 | 8.8 | 15.7 |
| Clinical Officer | 146 | 42.2 | 37.1 | 47.5 |
| Medical Doctor | 159 | 46.0 | 40.7 | 51.2 |

### Preferences and marginal WTT and WTW estimates

Overall, the preference weights for each attribute level had the expected signs (Figure 1 and Table 7). NHIF members preferred facilities where drugs were always available (β = 1.572; p-value=<0.001), with health workers that were respectful (β = 1.249; p-value=<0.001), with clinical officers (β = 0.478; p-value=<0.001) or medical doctors (β = 1.525; p-value=<0.001) for consultation and were always cleaner (β = 0.689; p-value=<0.001). NHIF members did not prefer facilities that were further away from their households (β = -0.082; p-value=<0.001) or having to wait longer at the facility until they could get a consultation (β = -0.195; p-value=<0.001). All standard deviations across all attributes were significant (p-value<0.001) except for distance and waiting time indicating the presence of inter-respondent heterogeneity in preferences.

**Figure 1:**
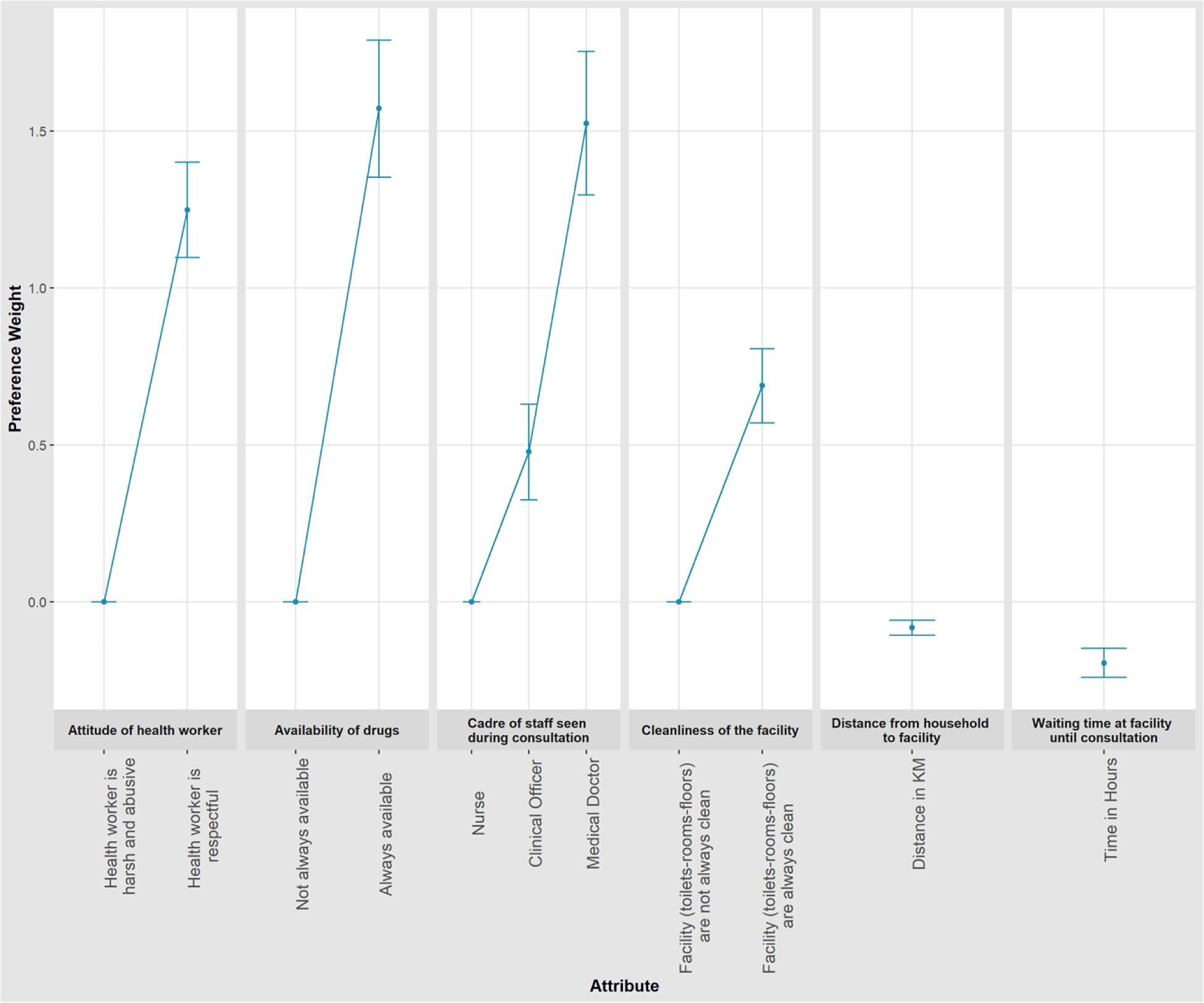
Mean preference weights from the panel MMNL model outputs. The change in utility associated with a change in the levels of each attribute is represented by the vertical distance between any two levels of the attribute. The utility of the base level was set at 0.0.

**Table 7:** Main effects panel MMNL model preference weights and marginal WTT and WTW estimates.

| Attributes | Preference estimates |  |  | Standard Deviation estimates |  |  |
| --- | --- | --- | --- | --- | --- | --- |
|  | Coefficient | 95% CI | P-value | Coefficient | 95% CI | P-value |
| **Availability of Drugs** |  |  |  |  |  |  |
| Not always available | Ref. (o) |  |  |  |  |  |
| Always available | 1.572 | 1.353 to 1.790 | <0.001 | 1.25 | 1.043 to 1.458 | <0.001 |
| **Distance from household to facility** | -0.082 | -0.106 to -0.058 | <0.001 | 0.002 | -0.01 to 0.013 | 0.768 |
| **Waiting time at the facility until consultation** | -0.195 | -0.241 to -0.148 | <0.001 | 0.018 | -0.025 to 0.060 | 0.413 |
| **Attitude of health workers** |  |  |  |  |  |  |
| Health worker is harsh and abusive | Ref. (o) |  |  |  |  |  |
| Health worker is respectful | 1.249 | 1.097 to 1.401 | <0.001 | 0.901 | 0.734 to 1.068 | <0.001 |
| **Cleanliness of the facility** |  |  |  |  |  |  |
| Facility (toilets-rooms-floors) are not always clean | Ref. (o) |  |  |  |  |  |
| Facility (toilets-rooms-floors) are always clean | 0.689 | 0.570 to 0.807 | <0.001 | 0.658 | 0.494 to 0.821 | <0.001 |
| **Cadre of health worker seen during a consultation** |  |  |  |  |  |  |
| Nurse | Ref. (o) |  |  |  |  |  |
| Clinical Officer | 0.478 | 0.325 to 0.630 | <0.001 | 0.608 | 0.417 to 0.799 | <0.001 |
| Medical Doctor | 1.525 | 1.296 to 1.753 | <0.001 | 0.837 | 0.594 to 1.079 | <0.001 |
| **Model fit statistics** |  |  |  |  |  |  |
| Log-likelihood (final) | -2728.892 |  |  |  |  |  |
| Number of Observations | 9648 |  |  |  |  |  |
| Number of decision-makers (n) | 402 |  |  |  |  |  |
| Draws (Halton) | 1000 |  |  |  |  |  |

Findings from the willingness to pay (WTP) space for the marginal willingness to accept travel (WTT) and willingness to accept waiting (WTW) at a facility highlighted key trade-offs among NHIF members (Table 8). First, on WTT, NHIF members were willing to travel up to 17.8 kilometres [(95% CI: 12.7 – 22.9), p-value<0.001] to a facility where drugs were always available, compared to a facility where they were not. Besides, NHIF members were willing to travel 14.6, 7.7, 5.7, and 17.7 kilometres to an NHIF-contracted outpatient facility where health workers were respectful, it was always clean and could be seen by a clinical officer and medical doctor for consultation compared to a facility where health workers were not respectful, not always clean and could see a nurse for consultation respectively. However, NHIF members were willing to accept a reduction of up to 2.0 Kilometres [(95% CI: -2.8 to -1.2), p-value<0.001] for an additional hour of waiting time at a facility until they got a consultation.

**Table 8:** Marginal willingness to travel (WTT) and willingness to wait (WTW) estimates in willingness to pay space.

| Attributes | Marginal WTT estimates in WTP space |  |  | Marginal WTW estimates in WTP space |  |  |
| --- | --- | --- | --- | --- | --- | --- |
|  | Coefficient | 95% CI | P-value | Coefficient | 95% CI | P-value |
| **Availability of Drugs** |  |  |  |  |  |  |
| Not always available | Ref. (o) |  |  | Ref. (o) |  |  |
| Always available | 17.839 | 12.744 to 22.933 | <0.001 | 8.885 | 6.376 to 11.394 | <0.001 |
| **Distance from household to facility** | WTT denominator |  |  | -0.497 | -0.680 to -0.314 | <0.001 |
| **Waiting time at the facility until consultation** | -2.03 | -2.816 to -1.244 | <0.001 | WTW denominator |  |  |
| **Attitude of health workers** |  |  |  |  |  |  |
| Health worker is harsh and abusive | Ref. (o) |  |  | Ref. (o) |  |  |
| Health worker is respectful | 14.617 | 10.728 to 18.506 | <0.001 | 7.207 | 4.975 to 9.439 | <0.001 |
| **Cleanliness of the facility** |  |  |  |  |  |  |
| Facility (toilets-rooms-floors) are not always clean | Ref. (o) |  |  | Ref. (o) |  |  |
| Facility (toilets-rooms-floors) are always clean | 7.739 | 5.182 to 10.295 | <0.001 | 3.804 | 2.612 to 4.996 | <0.001 |
| **Cadre of health worker seen during a consultation** |  |  |  |  |  |  |
| Nurse | Ref. (o) |  |  | Ref. (o) |  |  |
| Clinical Officer | 5.69 | 3.651 to 7.729 | <0.001 | 2.833 | 1.560 to 4.106 | <0.001 |
| Medical Doctor | 17.743 | 12.657 to 22.830 | <0.001 | 8.827 | 6.249 to 11.405 | <0.001 |
| **Model fit statistics** |  |  |  |  |  |  |
| Log-likelihood (final) | -2722.291 |  |  | -2718.378 |  |  |
| Number of Observations | 9648 |  |  | 9648 |  |  |
| Number of decision-makers (n) | 402 |  |  | 402 |  |  |
| Draws (Halton) | 1000 |  |  | 1000 |  |  |

Second, when waiting time was considered as the “monetary or price” attribute, similar trends were observed. For instance, NHIF members were willing to wait at a facility for up to 8.9, 8.8, 7.2, 3.8, and 2.8 hours as long as they would always get drugs, could be seen by a medical doctor during a consultation, health workers were respectful, the facility toilets, rooms, and floors were always cleaner and could be seen by a clinical officer during consultation respectively. However, all things constant, NHIF members were willing to accept a reduction in waiting time of up to 30 minutes [0.5 hours (95% CI: -0.7 to -0.3), p-value<0.001] if they had to travel to a facility that was 1 Kilometre further away from their households.

### Relative importance of the attributes

Table 9 and Figure 2 summarize the findings of the exploration of the most important attributes.The cadre of staff seen during a consultation was the most important attribute (0.277 [95% CI: 0.276 – 0.278]) followed by availability of drugs (0.273; 95% CI: 0.272 – 0.273), attitude of health worker (o.216 [95% CI: 0.216 – 0.217]), and cleanliness of the facility (0.123 [0.122 – 0.123]). Waiting time (0.053 [95% CI: 0.053 – 0.053]) and distance from the household to a facility (0.059 [95% CI: 0.058 – 0.059]) had the least importance scores.

**Figure 2:**
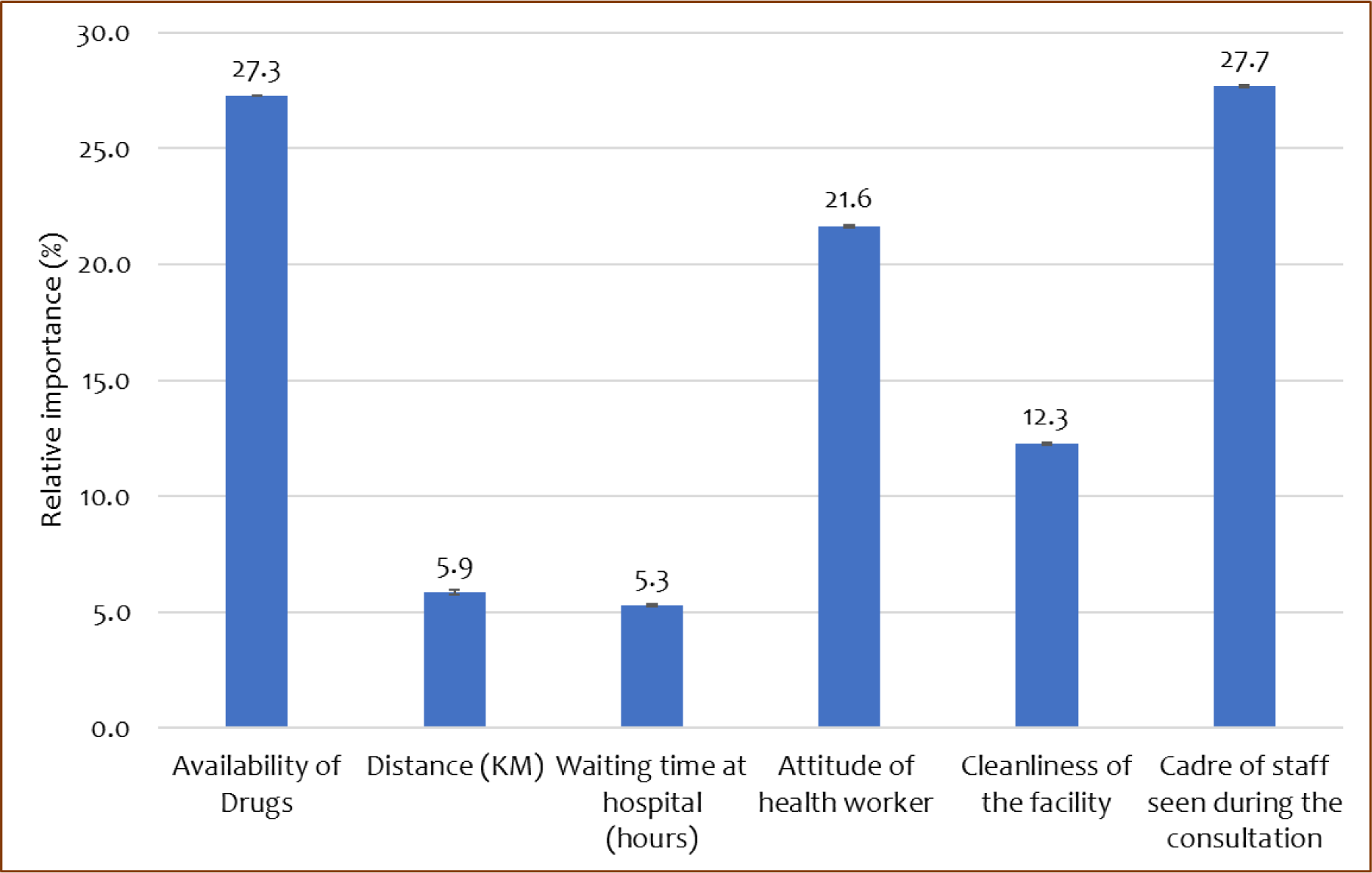
Relative importance of attributes in percentage.

**Table 9:** Relative importance of attributes.

| Attribute | Effect | Maximum Effect | Relative Importance |  |  |  |
| --- | --- | --- | --- | --- | --- | --- |
|  |  |  | Relative Importance | Standard error | 95% CI |  |
|  |  |  |  |  | Lower | Upper |
| Availability of Drugs | 1.101 | 1.100 | 0.273 | 0.00045 | 0.272 | 0.273 |
| Distance from household to the facility | -0.059 | 0.236 | 0.059 | 0.00028 | 0.058 | 0.059 |
| Waiting time at the facility for consultation | -0.143 | 0.214 | 0.053 | 0.00020 | 0.053 | 0.053 |
| Attitude of health worker | 0.874 | 0.873 | 0.216 | 0.00033 | 0.216 | 0.217 |
| Cleanliness of the facility | 0.493 | 0.495 | 0.123 | 0.00029 | 0.122 | 0.123 |
| Cadre of staff seen during the consultation | 1.119 | 1.118 | 0.277 | 0.00047 | 0.276 | 0.278 |
| Total |  | 4.037 |  |  |  |  |

### Unobservable preference heterogeneity

Figure 3 presents the findings from the three-class LC model (Supplementary File 2). The largest class (hereafter referred to as the ‘drug-focused’ class) had a membership probability of 63.4%. Preferences of participants most likely to be in this class were primarily driven by drugs being always available as opposed to drugs not being always available [β=1.55; p-value<0.001]. Besides, members in this class also had lower preferences for facilities that were further away from their households [β=-0.06; p-value<0.001] and had longer waiting times [β=-0.167; p-value<0.001].

**Figure 3:**
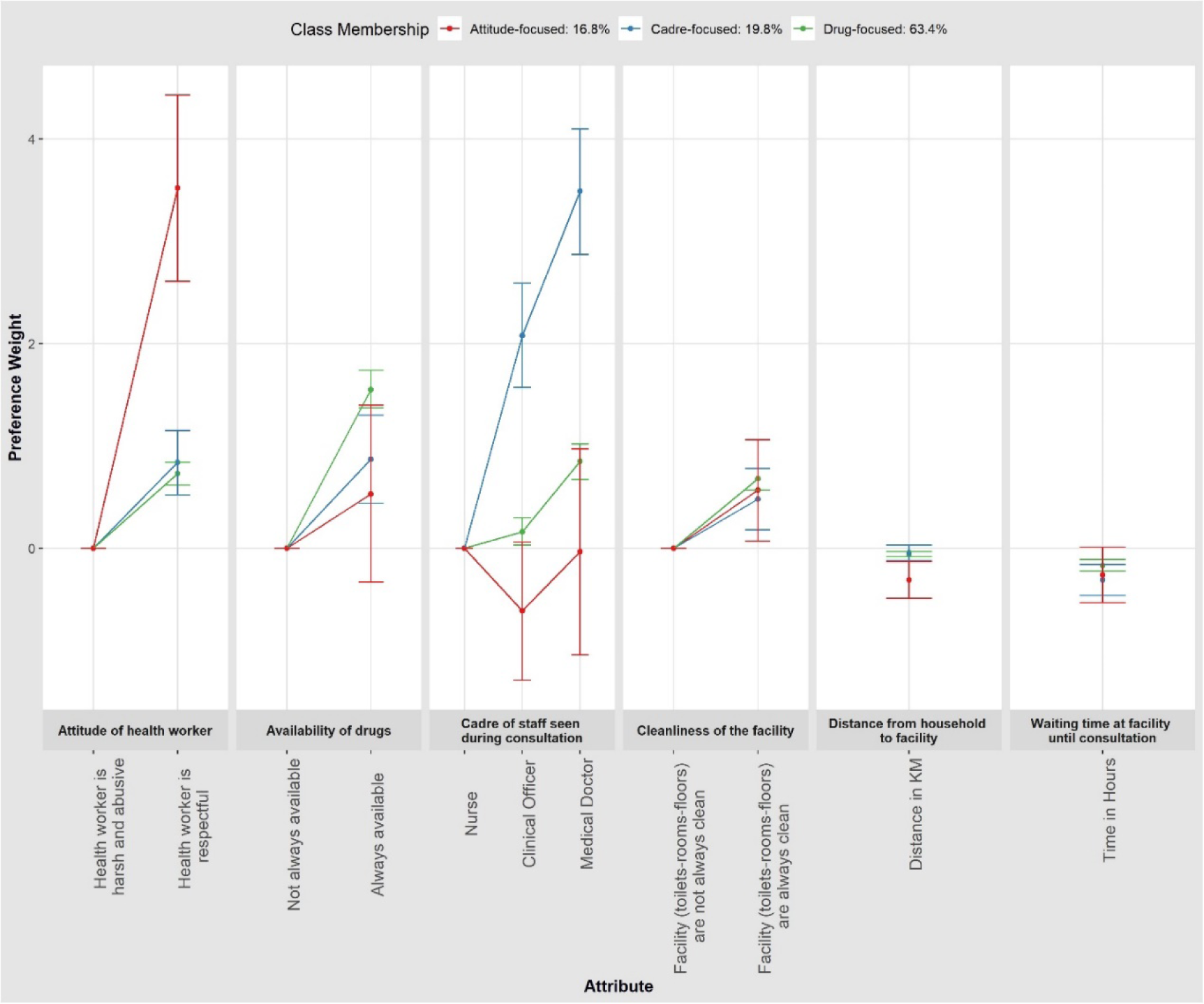
Latent Class Model results.

The second largest class (hereafter referred to as the ‘cadre-focused’ class) had a class membership probability of 19.8%. The preferences of members in this class were primarily driven by the cadre of staff seen during a consultation, particularly medical doctors [β=3.49; p-value<0.001] and clinical officers [β=2.08; p-value<0.001] as opposed to nurses, hence referred to here as the ‘cadre-focused’ class. Other preferences were similar to those described above.

The third and smallest class (hereafter referred to as the ‘attitude-focused’ class) had a membership probability of 16.8%. In this class, NHIF members’ choices were driven by the attitudes of health workers [β=3.52; p-value<0.001]. Interestingly, members of this class were averse to the cadre of staff seen during a consultation ((Medical Doctors [β=-0.03;p-value=0.950]); Clinical Officers [β=-0.615;p-value=0.073] even though this was not statistically significant.

## Discussion

Patient choice is increasingly gaining attention as an approach to enhancing provider competition and providing patient-centred care towards attaining health system goals. Understanding the preferences of users especially in low- and middle-income countries (LMICs) such as Kenya is crucial to informing decision-makers design of patient-centred care that aligns with UHC goals. This study examined the preferences of NHIF members for the attributes of NHIF-contracted outpatient facilities, marginal WTT and WTW, and preference heterogeneities. We found that, first, NHIF members preferred NHIF-contracted outpatient facilities where drugs were always available, were closer to their households, had shorter waiting times, had respectful health workers, and could see clinical officers or medical doctors during consultation. This is further strengthened by the findings from the relative importance of the attributes where the cadre of staff seen during a consultation, availability of drugs, and attitude of health workers were found to be the top three most important attributes. Second, NHIF members were willing to accept travelling to a further away facility or wait longer at a facility if the facility always had drugs, respectful health workers, and could be seen by a clinical officer or medical doctor. Third, our findings highlighted inter-respondent heterogeneity in NHIF members’ preferences and WTT and WTW parameters. Besides, we established three class memberships where choices in these classes were driven by the availability of drugs, the cadre of staff seen during a consultation, and the attitude of health workers. These findings can be explained.

NHIF members preferred facilities where drugs were always available. These findings are similar to those reported in other studies (9, 11, 51). Besides, the findings on the relative importance of attributes reflect the NHIF members’ focus on quality-related attributes which further highlights the importance of quality in patient-centred health systems (52). This is further corroborated by our WTT and WTW estimates where NHIF members indicated a willingness to accept travelling up to 17.8 kilometres from their households and wait up to 8.9 hours at an NHIF-contracted outpatient facility that always had drugs (Table 8Table 7). This could be explained by the contribution of drugs or medication costs to overall healthcare costs. For instance, studies in Kenya and other settings have highlighted drugs as the major contributor to healthcare costs (53–55). While ideally medicines should be provided for free to NHIF members seeking outpatient care, a recent study in Kenya did not find any significant evidence that NHIF provided financial protection to NHIF members with hypertension and diabetes (56). Even though that study was disease-specific, it highlights some nuances to the continued payment of out-of-pocket payments for services such as medication or drugs that should rather be free.

NHIF members preferred NHIF-contracted health facilities that were closer and had shorter waiting times. This finding corroborates evidence from other studies that have reported distance and waiting time as major factors influencing the choice of facilities (12, 34). Distance is crucial, especially in a setting like Kenya where transport costs were estimated to account for 31.4% of direct healthcare costs (53). Besides, evidence from another study in Kenya highlighted transport costs as the second largest contributor to the total direct diabetes healthcare costs (54). On the other hand, waiting time until consultation has been reported as the most used ‘structure’ attribute (12). In a Danish study, Pedersen et al. found waiting time until an appointment to be the most preferred attribute, more than the distance to the facility (57). In another study that examined patient preferences for facilities in Western Cape Province, Chiwire et al. also found waiting time as the second most important attribute (9). While gender differences were also reported in the Western Cape Province study, gender interaction with waiting time in our study did not reveal any significant difference between male and female NHIF members’ waiting time. However, this could be attributed to the high informality in employment (Table 5) and poverty levels in Kenya. Perhaps, NHIF members would want to resume their jobs in the informal settings given the lack of access to social protections often offered to formal sector workers such as sick leave days (58).

NHIF members preferred NHIF-contracted outpatient facilities with respectful health workers. These findings are similar to those reported in Tanzania where respondents preferred health facilities where doctors provided them with kind/respectful treatment (20). Similarly, Aridi et al. found women preferred health facilities that had kind and supportive health workers (19). This is in line with the quality of care standards where patients are entitled to receive treatment respectfully (59).

The cadre of staff seen during a consultation was also an important factor that influenced the choice of NHIF-contracted outpatient facilities among NHIF members. Most importantly, NHIF members preferred facilities where clinical officers or medical doctors would attend to them during consultation. Similar findings have been reported elsewhere. For instance, Caldow et al showed that respondents preferred to be attended to by a general practitioner (GP) than a practice nurse (60). While the preference for a specific cadre of staff may result from the perceived severity of a condition, clinical officers and medical doctors are perceived to be better skilled and knowledgeable in diagnosing illnesses as opposed to nurses (60).

### Implications for policy

Findings from this study offer several implications for policy in Kenya and other countries with similar settings. First, contracting outpatient facilities by purchasers such as the NHIF should prioritize facilities that always have drugs. For example, given the willingness to accept travelling and waiting at facilities, counties should prioritize equipping available facilities with required medication rather than building newer facilities that end up being ill-equipped with staff and commodities.

Second, interventions that aim to enhance the quality of care delivered across facilities should prioritize addressing waiting times and the attitudes of health workers. For instance, health workers could receive refresher training periodically to enhance their responsiveness and attitudes (particularly in treating patients with respect) when engaging with NHIF members and patients in general. These will then enhance user satisfaction whilst promoting the attainment of the responsiveness health system goal. Besides, there is a need to strengthen monitoring and accountability systems such as patient feedback mechanisms as levers for providing patient-centred care.

Third, given that the health workforce is a central health system building block (61), there is a need for the Kenyan government to deploy more health workers across the facilities particularly clinical officers and medical doctors. Besides, NHIF should strengthen its purchasing function to ensure the availability of required health workers across all the facilities it contracts.

### Study strengths and limitations

First, this is the first study that quantitatively examined the preference of NHIF members for attributes of NHIF-contracted outpatient facilities in Kenya. Given the role of NHIF as the main public health purchaser and the ‘vehicle’ of the UHC agenda in Kenya, these findings are crucial to providing evidence of who the NHIF should contract. Second, our study provides details of the trade-offs NHIF members were willing to make which can inform the tailoring of interventions such as employment of health workers, construction of facilities and equipping of facilities with essential medicines and supplies. Third, the use of the DCE in this study allowed the examination of NHIF members’ preferences in the absence of revealed preference data.

However, these findings should be interpreted in light of the following limitations. Methodologically, DCEs have been associated with hypothetical bias resulting from the difference between what people say they would do versus what they actually do (62, 63). While we aimed to assess the external validity (hypothetical bias) in this study and we collected data for respondents that had selected an NHIF-contracted outpatient facility, we could not fit models using the revealed preference data due to inadequate specification of the revealed preference choice variable. Future studies should assess this. Finally, the purposiveness in sampling NHIF members may have introduced a bias, however, we don’t anticipate this to have affected the findings.

## Conclusion

Findings from this study highlight the preferred NHIF-contracted outpatient health facility attributes and trade-offs NHIF members are willing to make. Consequently, there is a need for the NHIF, counties and health providers to prioritize these attributes and trade-offs when contracting providers, allocating resources, and providing services respectively. These attributes offer insights for the recently formed Social Health Authority (SHA) as the institution takes over the mandate of NHIF.

## Author Contribution

**Conceptualization:** Jacob Kazungu

**Formal Analysis:** Jacob Kazungu, Matthew Quaife

**Methodology:** Jacob Kazungu, Edwine Barasa, Justice Nonvignon, Mathew Quaife

**Visualization:** Jacob Kazungu

**Writing – original draft:** Jacob Kazungu

**Writing – review & editing:** Jacob Kazungu, Edwine Barasa, Justice Nonvignon, Matthew Quaife

**Supervision:** Edwine Barasa, Justice Nonvignon, Matthew Quaife

## Acknowledgments

This work was funded by Wellcome Trust Masters Fellowship grant number 212347 awarded to JK. The funders had no role in the study design, data analysis, decision to publish, drafting, or submission of the manuscript.

## DATA AVAILABILITY STATEMENT

The data that support the findings of this study are available on request from the corresponding author.

## References

1. MOH. Roadmap towards Universal Health Coverage in Kenya 2018–2022. 2018.

2. MOH. Kenya Health Policy 2014–2030: Towards attaining the highest standard of health. 2014.

3. HEFREP. The NHIF we want - Report of the Health Financing Reforms Expert Panel for the transformation and repositioning of National Hospital Insurance Fund as a strategic purchaser of health servises for the attainment of Universal Health Coverage by 2022. 2019.

4. Mbau R, Kabia E, Honda A, Hanson K, Barasa E. Examining purchasing reforms towards universal health coverage by the National Hospital Insurance Fund in Kenya. International journal for equity in health. 2020;19:1–18.

5. Honda A. What is strategic purchasing for health? 2014.

6. Ali S, Ronaldson S. Ordinal preference elicitation methods in health economics and health services research: using discrete choice experiments and ranking methods. British medical bulletin. 2012;103(1):21–44.

7. Hensher DA, Rose JM, Greene WH. Applied choice analysis: a primer: Cambridge university press; 2005.

8. Ryan M, Gerard K, Amaya-Amaya M. Using discrete choice experiments to value health and health care: Springer Science & Business Media; 2007.

9. Chiwire P, Beaudart C, Evers SM, Mahomed H, Hiligsmann M. Enhancing public participation in public health offerings: patient preferences for facilities in the western cape province using a discrete choice experiment. International Journal of Environmental Research and Public Health. 2022;19(1):590.

10. Dixon A, Robertson R, Appleby J, Burge P, Devlin NJ. Patient choice: how patients choose and how providers respond: King’s Fund; 2010.

11. Dündar C. Health-seeking behavior and medical facility choice in Samsun, Turkey. Health Policy. 2017;121(9):1015–9.

12. Kleij K-S, Tangermann U, Amelung VE, Krauth C. Patients’ preferences for primary health care–a systematic literature review of discrete choice experiments. BMC health services research. 2017;17(1):476.

13. Kuunibe N, Dary SK. Choice of healthcare providers among insured persons in Ghana. Research on Humanities and Social Sciences. 2012;2(10):88–97.

14. Lagarde M, Erens B, Mays N. Determinants of the choice of GP practice registration in England: evidence from a discrete choice experiment. Health Policy. 2015;119(4):427–36.

15. Lendado TA, Bitew S, Elias F, Samuel S, Assele DD, Asefa M. Effect of hospital attributes on patient preference among outpatient attendants in Wolaita Zone, Southern Ethiopia: discrete choice experiment study. BMC health services research. 2022;22(1):1–11.

16. Liu Y, Kong Q, de Bekker-Grob EW. Public preferences for health care facilities in rural China: a discrete choice experiment. Social Science & Medicine. 2019;237:112396.

17. Kruk ME, Rockers PC, Tornorlah Varpilah S, Macauley R. Population preferences for health care in liberia: insights for rebuilding a health system. Health services research. 2011;46(6pt2):2057–78.

18. Jayanthi T, Suresh S, Padmanaban P. Primary health centres: preferred option for birthing care in Tamilnadu, India, from users’ perspectives. Journal of health, population, and nutrition. 2015;33(1):177.

19. Oluoch-Aridi J, Adam MB, Wafula F, Kokwaro G. Understanding what women want: eliciting preference for delivery health facility in a rural subcounty in Kenya, a discrete choice experiment. BMJ open. 2020;10(12):e038865.

20. Larson E, Vail D, Mbaruku GM, Kimweri A, Freedman LP, Kruk ME. Moving toward patient-centered care in Africa: a discrete choice experiment of preferences for delivery care among 3,003 Tanzanian women. PloS one. 2015;10(8):e0135621.

21. Kenya LO. The constitution of Kenya: 2010: Chief Registrar of the Judiciary; 2013.

22. Kazungu J, Mbithi L, Onyes U, Nwaononiwu E, Marangu M, Mamo A, et al. Changing the game in purchasing health services: findings from a provider-purchaser engagement in Kenya. 2022.

23. KNBS and ICF. Kenya Demographic and Health Survey 2022. Key Indicators Report. Nairobi, Kenya and Rockville, Maryland, USA: KNBS and ICF; 2023.

24. Barasa E, Kazungu J, Nguhiu P, Ravishankar N. Examining the level and inequality in health insurance coverage in 36 sub-Saharan African countries. BMJ Global Health. 2021;6(4):e004712.

25. Mbau R, Kabia E, Honda A, Hanson K, Barasa E. Examining purchasing reforms towards universal health coverage by the National Hospital Insurance Fund in Kenya. International journal for equity in health. 2020;19(1):19.

26. Barasa E, Rogo K, Mwaura N, Chuma J. Kenya National Hospital Insurance Fund Reforms: Implications and Lessons for Universal Health Coverage. Health Systems & Reform. 2018;4(4):346–61.

27. Hauber AB, González JM, Groothuis-Oudshoorn CG, Prior T, Marshall DA, Cunningham C, et al. Statistical methods for the analysis of discrete choice experiments: a report of the ISPOR conjoint analysis good research practices task force. Value in health. 2016;19(4):300–15.

28. Bridges JF, Hauber AB, Marshall D, Lloyd A, Prosser LA, Regier DA, et al. Conjoint analysis applications in health—a checklist: a report of the ISPOR Good Research Practices for Conjoint Analysis Task Force. Value in health. 2011;14(4):403–13.

29. Obadha M, Chuma J, Kazungu J, Abiiro GA, Beck MJ, Barasa E. Preferences of healthcare providers for capitation payment in Kenya: a discrete choice experiment. Health Policy and Planning. 2020;35(7):842–54.

30. Abuya T, Mwanga D, Obadha M, Ndwiga C, Odwe G, Kavoo D, et al. Incentive preferences for community health volunteers in Kenya: findings from a discrete choice experiment. BMJ open. 2021;11(7):e048059.

31. Oluoch-Aridi J, Adam MB, Wafula F, K’okwaro G. Eliciting women’s preferences for place of child birth at a peri-urban setting in Nairobi, Kenya: A discrete choice experiment. Plos one. 2020;15(12):e0242149.

32. Takemura T, Kielmann K, Blaauw D. Job preferences among clinical officers in public sector facilities in rural Kenya: a discrete choice experiment. Human resources for health. 2016;14(1):1–10.

33. Helter TM, Boehler CEH. Developing attributes for discrete choice experiments in health: a systematic literature review and case study of alcohol misuse interventions. Journal of substance use. 2016;21(6):662–8.

34. Kazungu J, Quaife M, Nonvignon J, Barasa E. What influences patients’ choice of health facilities and does it enhance provider competition in low- and middle-income countries? A scooping review [Unpublished manuscript]. 2024.

35. Kazungu J, Nonvignon J, Quaife M, Barasa E. Assessing the choice of National Health Insurance Fund contracted outpatient facilities in Kenya: A qualitative study. Int J Health Plann Manage. 2023.

36. Choice Metrics. Ngene Verson 1.3. 0. Sydney, New South Wales: ChoiceMetrics Pty Ltd. 2021.

37. Watson V, Becker F, de Bekker-Grob E. Discrete choice experiment response rates: A meta-analysis. Health economics. 2017;26(6):810–7.

38. de Bekker-Grob EW, Ryan M, Gerard K. Discrete choice experiments in health economics: a review of the literature. Health economics. 2012;21(2):145–72.

39. Clark MD, Determann D, Petrou S, Moro D, de Bekker-Grob EW. Discrete choice experiments in health economics: a review of the literature. Pharmacoeconomics. 2014;32:883–902.

40. Aoyama H. A study of stratified random sampling. Ann Inst Stat Math. 1954;6(1):1–36.

41. Kenya Master Health facility List [Internet]. 2021 [cited 25/04/2021]. Available from: http://kmhfl.health.go.ke/#/facility_filter.

42. Outpatient services: List of Medical Facilities Offering National Scheme [Internet]. 2019 [cited 13/11/2019]. Available from: http://www.nhif.or.ke/healthinsurance/outpatientServices.

43. Chorus C, van Cranenburgh S, Dekker T. Random regret minimization for consumer choice modeling: Assessment of empirical evidence. Journal of Business Research. 2014;67(11):2428–36.

44. Lancaster KJ. A new approach to consumer theory. Journal of political economy. 1966;74(2):132–57.

45. Lancsar E, Fiebig DG, Hole AR. Discrete choice experiments: a guide to model specification, estimation and software. Pharmacoeconomics. 2017;35:697–716.

46. Maaya L, Meulders M, Surmont N, Vandebroek M. Effect of environmental and altruistic attitudes on willingness-to-pay for organic and fair trade coffee in Flanders. Sustainability. 2018;10(12):4496.

47. Hole AR, Kolstad JR. Mixed logit estimation of willingness to pay distributions: a comparison of models in preference and WTP space using data from a health-related choice experiment. Empirical Economics. 2012;42:445–69.

48. StataCorp. Stata Statistical Software: Release 16.1. 2021.

49. Hole AR, editor Mixed logit modeling in Stata--an overview. United Kingdom Stata Users’ Group Meetings 2013; 2013: Stata Users Group.

50. Walsh DA, Boeri M, Abraham L, Atkinson J, Bushmakin AG, Cappelleri JC, et al. Exploring patient preference heterogeneity for pharmacological treatments for chronic pain: a latent class analysis. European Journal of Pain. 2022;26(3):648–67.

51. Honda A, Ryan M, van Niekerk R, McIntyre D. Improving the public health sector in South Africa: eliciting public preferences using a discrete choice experiment. Health policy and planning. 2015;30(5):600–11.

52. Kruk ME, Pate M. The Lancet global health Commission on high quality health systems 1 year on: progress on a global imperative. The Lancet global health. 2020;8(1):e30–e2.

53. Barasa EW, Maina T, Ravishankar N. Assessing the impoverishing effects, and factors associated with the incidence of catastrophic health care payments in Kenya. International journal for equity in health. 2017;16(1):31.

54. Oyando R, Njoroge M, Nguhiu P, Sigilai A, Kirui F, Mbui J, et al. Patient costs of diabetes mellitus care in public health care facilities in Kenya. The International journal of health planning and management. 2020;35(1):290–308.

55. Kazungu J, Meyer CL, Sargsyan KG, Qaiser S, Chukwuma A. The burden of catastrophic and impoverishing health expenditure in Armenia: An analysis of Integrated Living Conditions Surveys, 2014–2018. PLOS Global Public Health. 2022;2(10):e0000494.

56. Oyando R, Were V, Koros H, Mugo R, Kamano J, Etyang A, et al. Evaluating the effectiveness of the National Health Insurance Fund in providing financial protection to households with hypertension and diabetes patients in Kenya. International journal for equity in health. 2023;22(1):107.

57. Pedersen LB, Kjær T, Kragstrup J, Gyrd-Hansen D. Do general practitioners know patients’ preferences? An empirical study on the agency relationship at an aggregate level using a discrete choice experiment. Value in Health. 2012;15(3):514–23.

58. Oladosu AO, Khai TS, Asaduzzaman M. Factors affecting access to healthcare for young people in the informal sector in developing countries: a systematic review. Frontiers in Public Health. 2023;11:1168577.

59. WHO. Standards for improving quality of maternal and newborn carein health facilities. 2016.

60. Caldow J, Bond C, Ryan M, Campbell NC, Miguel FS, Kiger A, et al. Treatment of minor illness in primary care: a national survey of patient satisfaction, attitudes and preferences regarding a wider nursing role. Health Expectations. 2007;10(1):30–45.

61. Organization WH. Everybody’s business--strengthening health systems to improve health outcomes: WHO’s framework for action. 2007.

62. Haghani M, Bliemer MC, Rose JM, Oppewal H, Lancsar E. Hypothetical bias in stated choice experiments: Part II. Conceptualisation of external validity, sources and explanations of bias and effectiveness of mitigation methods. Journal of choice modelling. 2021;41:100322.

63. Quaife M, Terris-Prestholt F, Di Tanna GL, Vickerman P. How well do discrete choice experiments predict health choices? A systematic review and meta-analysis of external validity. The European journal of health economics. 2018;19(8):1053–66.

